# Characteristics of a tattooed population and a possible role of tattoos as a risk factor for chronic diseases: Results from the LIFE-Adult-Study

**DOI:** 10.1101/2025.01.31.25321465

**Authors:** Loryn E. Theune, Narges Ghoreishi, Christine Müller-Graf, Christoph Engel, Kerstin Wirkner, Ronny Baber, Peter Laux, Andreas Luch, Michael Giulbudagian

## Abstract

Tattoos and permanent make-up (PMU) gain increasing popularity among the general population. There are indications that pigments or their fragments may translocate within the body, however knowledge about possible systemic adverse effects related to tattoos is very limited. We investigated the prevalence of systemic chronic health effects including cardiologic diseases, cancer and liver toxicity and their relationship with the presence and characteristics of tattoos and PMU as part of the LIFE-Adult-study, a population-based cohort study. The control group without tattoos was from the same cohort and adjusted for confounders based on age, sex, smoking-status, body mass index, alcohol consumption and socioeconomic status. Of 4,248 participants, 7.4% (n=320) had either a tattoo (4.7%; n=199) or a permanent make-up (3.1%; n=135), or both (n=14). Thereof 5% (16/320) reported medical complications in relation to their tattoos or PMUs. Regarding systemic chronic health effects, increased odds ratios among the tattooed population were found for cardiologic diseases and liver toxicity. For non-melanoma skin cancer, results from the tattoo questionnaire gave no indication for any tumour development at the site of the tattoo. Our results demonstrate an increased risk for cardiologic and liver effects in participants bearing a tattoo. Strong differences in the observed effects between men and women point on the presence of a sex-specific bias. Even if no statistically robust conclusion can be drawn due to the small sample size, the results suggest that cardiologic outcomes and liver toxicity in tattooed individuals should be investigated further using prospective designs in larger cohorts.

## Introduction

Tattoos have become increasingly popular particularly among younger people. In Europe, the prevalence of tattoos and permanent make-up (PMU) is estimated to be 10-20% among the general population, steadily increasing over the past decades [1–6]. In the United states, tattooing is even more popular with general prevalence of 31% and among the younger population of around 50% [7].

For both, tattoos and PMUs, coloured inks are injected into the dermis with the help of needles. The inks may contain pigments of organic or inorganic nature [8]. Most common organic pigments responsible for the colour brilliance are based on azo, quinacridone or Cu-phthalocyanine structures, while inorganic pigments originate from iron-oxides, carbon black or titanium dioxide [9]. Despite the increasing popularity of tattoos, knowledge about the biokinetics of pigments and other ink components is still limited. A fraction of colouring pigments remains at the injection site through uptake into tissue-resident macrophages and persist at their location via a “capture-release-recapture” mechanism assuring their (life-)long visibility [10–12]. Nevertheless, after injection the dermal pigment concentration decreases during wound healing and over the years, indicative for constant degradation and/or translocation processes ongoing [13]. Studies could show that pigments as well as other ink ingredients are translocated via lymph or blood vessels by migrating cells e.g., to local lymph nodes [13–16]. In addition, in animal studies after intradermal or subcutaneous injections of nanoparticles, these were detected in organs such as liver, lungs or kidney [16–18].

Amongst the known health risks, at least one third of chronic intolerance reactions to tattoos are allergic reactions [17, 19–25]. While often attributed to red pigments through temporal or spatial correlation, the causing allergen remains in most cases unidentified. Inflammatory reactions such as foreign-body granulomas (sarcoidosis) often associated with black tattoos are seen in clinics [23, 25]. Notably, along with localised reactions, extracutaneous involvement including uveitis and systemic sarcoidosis has been reported, which may indicate a systemic occurrence of pigment particles or fragments thereof [26, 27]. Tattoo related infective endocarditis is reported in the literature [28]. However, due to limited knowledge on the distribution of pigments, chemical and biological contaminants, along with the lacking prospective epidemiological studies, well-founded knowledge about possible adverse systemic health effects of tattoos is non-existent to date.

The present study, describes the occurrence and characteristics of tattoos and PMUs among an adult cohort in Germany and aims to investigate the role of tattoos in three health related outcomes, namely cardiologic diseases, liver toxicity and non-melanoma skin cancer.

## Methods

### Data Source and study participants

The Leipzig Research Centre for Civilization Diseases (LIFE) is conducting the LIFE-Adult-study, a population-based cohort study with the aim to investigate prevalence, early onset markers, genetic predisposition, and the role of lifestyle factors with relevance for major civilization diseases. From 2011 to 2014, the study recruited 10,000 randomly selected adult participants from Leipzig, Germany. In the baseline examination, all participants underwent an extensive core assessment programme including structured interviews/questionnaires, physical examinations, and bio specimen collection [29]. The first follow-up included questionnaires and a physical examination of selected participants [30]. The recruitment period for this study started on 11.06.2018 and ended on 11.12.2020.

The responsible ethics board at the Medical Faculty of the University of Leipzig approved the study (reference: LIFE Adult - 263-2009-14122009). All participants provided written informed consent to participate prior to participation.

### Exposure (Tattoo and PMU)

For the present study, a separate tattoo-specific questionnaire was (i) sent out to participants who agreed in the follow-up to be contacted again or (ii) issued upon physical follow-up examination. This questionnaire addressed the prevalence, size, location and colour of the tattoo and PMU as well as the occurrence of related medical complications (original questionnaire in German, see Fig. S1).

### Outcome

Serum levels of N-terminal pro-brain natriuretic peptide (NT-proBNP), as marker of cardiac insufficiency, and the enzymes alanine aminotransaminase (ALT) and aspartate aminotransaminase (AST), as well as the γ-glutamyltransferase (GGT), as biomarkers for liver toxicity, were obtained from blood samples at baseline.

Increased liver values indicative of hepatocellular damage (liver toxicity) was defined as increased level of ALT alone, as well as combinations of increased levels of ALT, AST and GGT, or AST and GGT [31–33]. This is because AST and ALT are both enzymes associated with liver parenchymal cells, which are found increased in the systemic circulation upon liver cell damage. While ALT is found predominantly in the liver with only minor quantities in other organs, AST is also present in other parts of the body in higher quantities such as heart (cardiac muscle) and skeletal muscles. As such AST is less liver specific and as well found elevated upon muscle damage or diseases related to cell damage such as myocardial infarcts, acute pancreatitis, acute hemolytic anemia, severe burns, acute renal disease, or musculoskeletal diseases. GGT is a liver and bile-duct specific amino acid transferase and elevated serum concentration is mainly correlated to hepatobiliary diseases and excessive alcohol or drug consumption. GGT has a low diagnostic specificity for liver diseases if it is the only enzyme elevated.

Occurrence, date and type of cancer, together with occurrence and date of myocardial infarction (MI), heart failure (HF), and any medical complication issues assumed to be related to a tattoo were obtained from questionnaire at baseline and follow-ups. The MI and HF outcomes were combined to a composite cardiovascular disease outcome.

### Confounders

We drew a directed acyclic graph (DAG) for the effect of tattoo on health related outcomes (Fig. S2). We also visualized our analysis based on the information available to control for confounding (Fig. S3). The sociodemographic and other baseline characteristics were obtained from the LIFE baseline investigation including sex, age, smoking status (non-, former or current smoker), body mass index (category 1-4), socioeconomic status (1 low-3 high), and alcohol consumption (gram per day). Liver toxicity (ALT, AST and GGT) as well as levels of NT-proBNP as marker of cardiac insufficiency were determined on the basis of laboratory values. Information on cardiovascular conditions, tumours, tobacco smoking, body mass index, alcohol consumption, and sociodemographic status are based on self-reports.

### Statistical Analyses

For the analysis, we matched the participants based on their tattoo status. We performed a full matching including all the confounding mentioned above to estimate the ATT (average effect of the treatment) in the treated population (i.e., average effect of tattoo/PMU). The full matching allows to use all the data in the analysis and avoid valuable participant data loss.

After matching, we used a weighted logistic regression to calculate the effect of the above mentioned health outcomes. We performed logistic regression on unmatched data to compare the results with the matched. The unmatched regression was adjusted for the confounders based on our directed acyclic graph (Fig. S2 and S3). Where necessary, we included an interaction term in the regression model. Statistical analysis was executed using R. For matching we used R package MatchIt [34, 35]. Graphics are drawn with R and Prism (GraphPad Prism Version 10.1.2 (324)).

For the outcomes with very low prevalence in the tattooed/PMU participants, the data was analysed descriptively.

### Sensitivity analysis

Since the study population is in East Germany and some participants were tattooed a long time before the outcome measurement, we performed a sensitivity analysis to see if there is an effect difference between the tattoos that took place before and after 1989 (reunification year of Germany). We assume that the composition of tattoo inks was different in GDR times and therefore differing effects are likely.

## Results

Fig shows the process of implementing the inclusion and exclusion criteria on the original LIFE-Adult cohort. Table 1 summarises the characteristics of the study sample and the sample size for each of the analyses. Among the tattooed/PMU participants the prevalence of non-melanoma skin cancer was very low. Hence, no analysis was done for this outcome.

**Table 1.** Characteristics of the total study sample.

| Characteristic | Overall, N = 4,295<sup>a</sup> | no-Tattoo, N = 3,975<sup>a</sup> | Tattoo/PMU, N = 320<sup>a</sup> | LIFE-Cohort, N = 10,000<sup>a</sup> |
| --- | --- | --- | --- | --- |
| **Gender** |  |  |  |  |
| Women | 2,241 (52%) | 2,020 (51%) | 221 (69%) | 5,234 (52%) |
| Men | 2,054 (48%) | 1,955 (49%) | 99 (31%) | 4,766 (48%) |
| **Age** | 69 (58, 78) | 70 (59, 78) | 63 (54, 72) | 68 (58, 78) |
| **Age categorized** |  |  |  |  |
| Elderly (> 60) | 2,941 (68%) | 2,769 (70%) | 172 (54%) | 6,684 (67%) |
| Adult (40-60) | 1,242 (29%) | 1,110 (28%) | 132 (41%) | 3,053 (31%) |
| Young (20-40) | 112 (2.6%) | 96 (2.4%) | 16 (5.0%) | 263 (2.6%) |
| **Body Mass Index (BMI)** | 26.4 (23.9, 29.7) | 26.5 (24.0, 29.7) | 25.2 (23.1, 28.9) | 26.6 (23.9, 30.0) |
| **BMI categorized** |  |  |  |  |
| Underweight | 17 (0.4%) | 15 (0.4%) | 2 (0.6%) | 58 (0.6%) |
| Normal | 1,525 (36%) | 1,374 (35%) | 151 (47%) | 3,413 (34%) |
| Overweight | 1,764 (41%) | 1,660 (42%) | 104 (33%) | 4,020 (40%) |
| Obese | 984 (23%) | 921 (23%) | 63 (20%) | 2,473 (25%) |
| Unknown | 5 (0.1%) | 5 (0.1%) | 0 (0%) | 36 (0.4%) |
| **Socioeconomic status (SES)** |  |  |  |  |
| Low | 631 (15%) | 573 (14%) | 58 (18%) | 1,995 (20%) |
| Medium | 2,612 (61%) | 2,406 (61%) | 206 (64%) | 5,994 (60%) |
| High | 1,041 (24%) | 986 (25%) | 55 (17%) | 1,974 (20%) |
| Unknown | 11 (0.3%) | 10 (0.3%) | 1 (0.3%) | 37 (0.4%) |
| **Alcohol consumption (g/day)** | 5 (1, 16) | 5 (1, 16) | 4 (1, 14) | 4 (1, 15) |
| **Alcohol consumption (categorized – g/day)<sup>b</sup>** |  |  |  |  |
| [0-10) | 2,478 (58%) | 2,282 (57%) | 196 (61%) | 5,746 (57%) |
| [10-30) | 817 (19%) | 752 (19%) | 65 (20%) | 1,712 (17%) |
| [30-50) | 345 (8.0%) | 326 (8.2%) | 19 (5.9%) | 748 (7.5%) |
| [50-193] | 178 (4.1%) | 165 (4.2%) | 13 (4.1%) | 427 (4.3%) |
| Unknown | 477 (11%) | 450 (11%) | 27 (8.4%) | 1,367 (14%) |
| **Smoking status** |  |  |  |  |
| Non | 2,257 (53%) | 2,141 (54%) | 116 (36%) | 4,680 (47%) |
| Former | 1,245 (29%) | 1,132 (28%) | 113 (35%) | 2,805 (28%) |
| Current | 707 (16%) | 621 (16%) | 86 (27%) | 2,105 (21%) |
| Unknown | 86 (2.0%) | 81 (2.0%) | 5 (1.6%) | 410 (4.1%) |
| **PMU** |  |  |  |  |
| No | 4,069 (96%) | 3,928 (100%) | 141 (46%) | - |
| Yes | 135 (3.2%) | 0 (0%) | 135 (48%) | - |
| Unknown | 39 (0.9%) | 0 (0%) | 39 (13%) | - |

<sup>a</sup> n (%) or (median interquartile range (IQR));
<sup>b</sup> ][ The number is inside the range; )( The number is not included in the range.

### Characteristics of tattooed/PMUed sub-cohort

In total, 4,308 participants responded to the tattoo questionnaire (response rate 86%; 5,000 questionnaires sent out). Of all respondents, 199/4,308 participants reported to be tattooed (4.7%) and 135/4,308 (3.1%) to have a PMU, and 14 (0.3%) stated to have both. Out of 199 tattooed individuals 159 received their tattoo prior to the baseline examination. Less than half (50 out of 135) of the participants with PMU received it prior to baseline examination. The incidence of a tattoo or PMU among the present cohort population was 7.5%. For the assessment of chronic effects 10 participants with an unreported date of receiving their tattoo/PMU were excluded from the analysis.

Among tattooed persons, 97 (48%) were men and 104 (52%) were women while for PMU the majority (129, 96%) were women (Fig. 2a). Median age for getting a tattoo was 30 years (IQR 19 years), while men (26 years, IQR 18 years) got their tattoo at a younger age than women (34 years, IQR 27 years) (Fig. 2b). In contrast, median age for getting a PMU was much higher with 59 years (IQR 16 years). The total percentage of participants having their tattoo/PMU longer than 10 years is higher for tattoos (79%) than for PMUs (39%) (Fig. 2c). The characteristics of the tattoo/PMU sub-cohort at baseline and follow-up is shown in Table 1.

**Fig 1.**
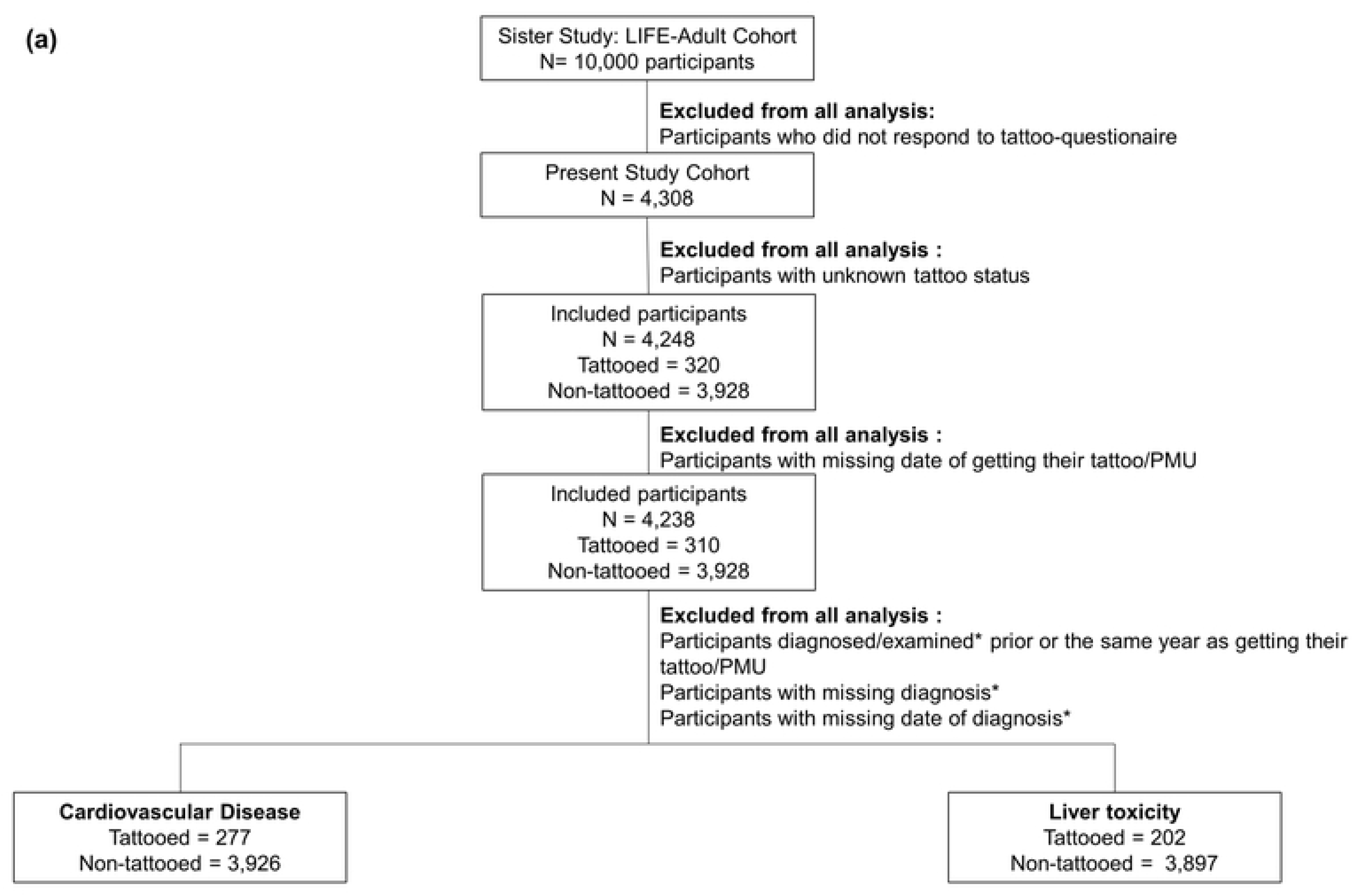
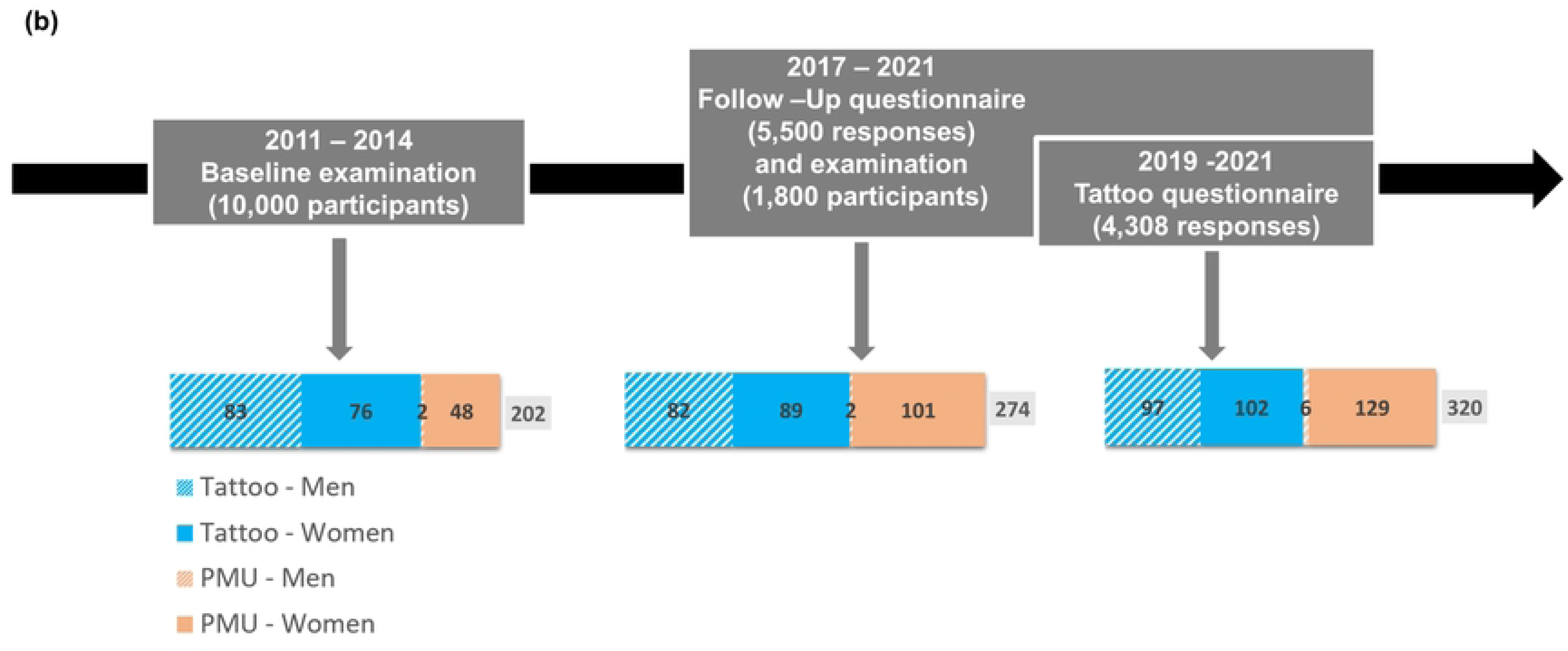
(a) Process of implementing the inclusion and exclusion criteria on the original LIFE cohort (b) Examination time frames and number of participants having a tattoo or PMU and overall number of participants with tattoo/PMU (grey, excluding double counting of participants having both tattoo and PMU) prior to a respective examination. *Diagnosis is referred to self-reported data in the questionnaire, while examination is referred to laboratory analysis of blood samples.

**Fig 2.**
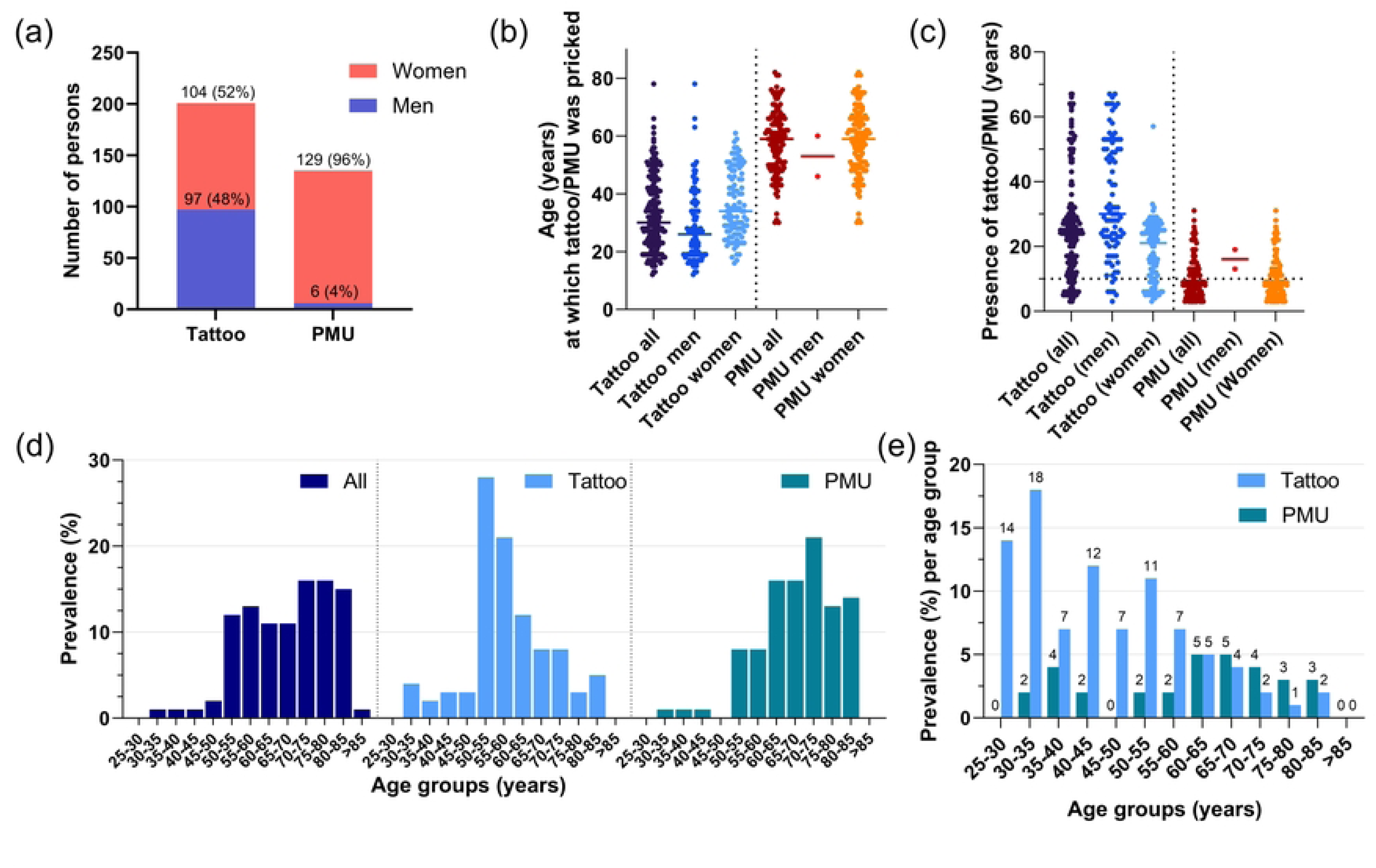
Sex and age distribution of the tattooed population. (a) Sex distribution among tattooed and PMUed participants, (b) Age at which tattoo or PMU was pricked depending on sex, (c) Presence of tattoo/PMU in years, (d) Age distribution among different subgroups (overall cohort, tattooed persons, and persons with PMU), (e) Prevalence of a tattoo and PMU among each age group separately (depicts the prevalence in relation to the size of the sup-cohort). It is to note that in very young age groups, particularly in the 25-30 years sub-cohort, the overall number of tattooed participants is low (25-30 years overall: 7 participants; 1 tattooed; 30-35 years: 49 participants overall, 9 tattooed).

While the overall LIFE cohort population has a right-skewed age distribution (median 68.5 years, IQR 20 years), the tattoo/PMU sub-cohort is younger with median of 57 years (IQR 13.5 years) (Fig. 2d).

Median tattoo size was 125 cm², the IQR was 250 cm²; 90^th^ and 95^th^ percentiles are 760 cm² and 1053 cm², respectively (Fig. 3, Fig. S4). All PMUs were in the face and except for one, all were reported to be small or medium sized. Larger tattoos were located on larger body parts, while small tattoos were distributed in all body parts. Location distributions were similar among men and women.

**Fig 3.**
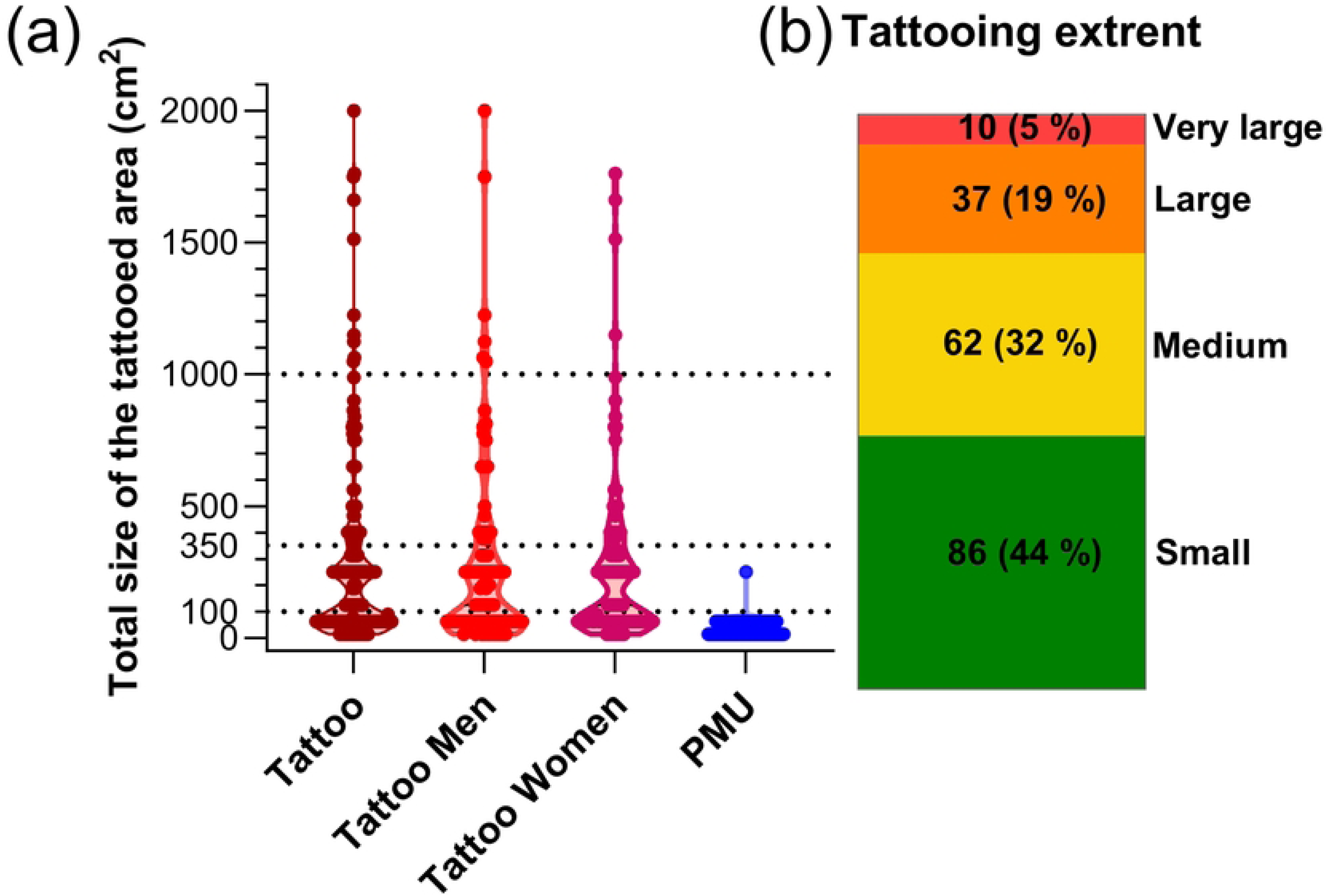
Size of the tattoos. (a) Individual sums of tattoo area (n=195) and size of PMUs (n=129). The area of the tattooed skin surface was calculated as a square of the given sizes (estimating the size as length of one side of the square). (b) Distribution of tattooing extent. Tattooing extent was estimated by calculating the individual sum of tattooed area from different body parts per participant (Fig. 4a) and categorized into four groups: small (indiv. sum <100 cm²), medium (indiv. sum 100 - 350 cm²), large (indiv. sum > 350 – 1000 cm²) and very large (>1000 cm²).

Black was the most common colour for both, tattoos 32.4% (126/389) and PMU 40.9% (65/159) (Fig. 4, Fig. S5). In tattoos, the prevalence of other colours such as blue, red, green, yellow, etc. was pronounced with similar distribution amongst men and women, while for PMUs only brown (40.9%, 69/159) and red (7.5 %, 12/159) were common. 38% (75/201) of tattooed participants were tattooed with more than one colour, 23% (46/201) with more than two colours. More than half (56%, 76/135) of the participants with a PMU stated that their PMU(s) have more than one colour, but only a minority indicated more than two colours (2%, 3/135).

**Fig 4.**
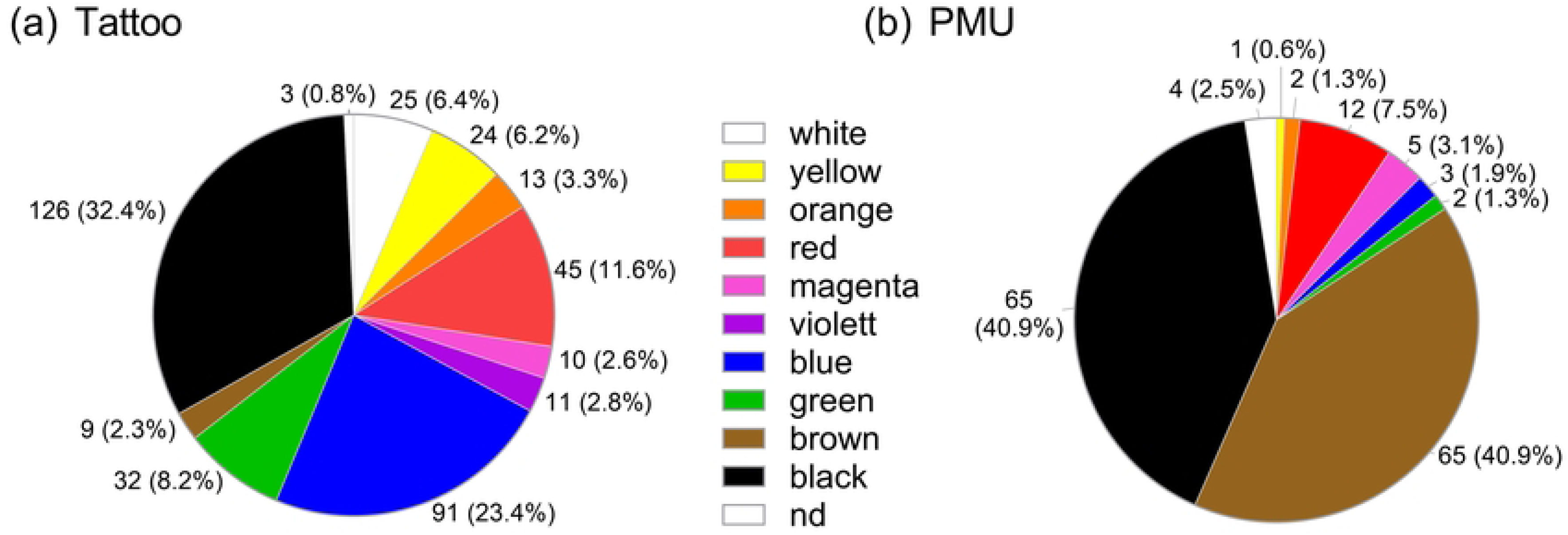
Prevalence of different colours in (a) tattoos or (b) PMUs.

The shares of current and former smokers among tattooed/PMUed people were generally higher than in the overall LIFE cohort with 29 % vs 22% current and 37% vs 29% former smokers at basis examination, respectively. Among women having a tattoo/PMU, the share of non-smokers (never smoked) is much higher 41% vs 21% and for current smokers significant lower 18% vs 42% than for men.

Median BMI at the basic examination was slightly lower (25.2 vs 26.6) among the tattooed/PMUed sub-cohort in comparison to the overall LIFE cohort. While among tattooed/PMUed sub-cohort around half of the population was in the normal BMI range (category 2) majority of the whole LIFE cohort had overweight (40%, BMI category 3). The fraction of women with overweight (28% vs 39%) an obesity (15% vs 27%) was lower than for men.

### Medical issues related to tattoos (self-report)

Of all tattooed/PMUed participants, 5% (16/320) reported on medical complications related to their tattoo or PMU (Table S1). Among them 5 had a tattoo, 9 had a PMU, and two had both.

Frequently named complications were pain, itchiness or swelling (10/320, 3.1%; 8/10 PMU, 2/10 tattoo). The tattooing extent among this group was mostly small and 7/10 of cases had a tattoo of black colour.

Other complications included herpes outbreak after receiving a PMU on the lips (n= 3, 0.9%), allergic tattoo/PMU reactions (n=2; 0.6%), scar formation (n=1, 0.3%) and a seldom and slight prickling at the tattoo site (n=1, 0.3%).

Thirty-two (10%) tattooed/PMUed participants (15 tattooed, 18 PMUed, 1 both) indicated to experience skin irritation through contact with metallic buttons, jewellery or similar items (indication for nickel allergy). However, only for 2 persons skin reactions occurred for the first time after receiving the tattoo or PMU. In both cases, the participants did not indicate a tattoo related medical issue.

### Cardiovascular diseases

The prevalence of myocardial infarction (MI) was 2.5% (7/277) in the tattooed/PMUed participants and 3% (120/3,926) in the non-tattooed participants (Table S2). Median age at first MI is higher for tattooed/PMUed (60 years, IQR 5 years) than for non-tattooed (54.5 years, IQR: 9 years). The time span between the occurrence of first MI and obtaining the tattoo was in average more than 30 years and minimum of 8 years. Information on cardiovascular diseases is based on self-reporting and hence not validated.

Overall, 4.0% (11/277) of the tattooed/PMUed participants and 4.1% (163/3,926) of the non-tattooed participants reported to be diagnosed with Heart failure (HF) (Table S2). Tattooing extent was small (4/9) and medium (5/9) only. With the exception of two cases, all participants have also experienced other cardiologic complications such as high blood pressure, atrial fibrillations, tachycardia and cardiac arrythmia amongst others.

In the sensitivity analysis we noticed a different effect among men and women and as a result we added an interaction term between tattoo/PMU status and sex in our regression model. Considering both cardiovascular outcomes (MI and HF) in the matched data the odds ratio of having a cardiovascular outcome for participants with tattoo was higher compared to non-tattooed participants (OR = 1.48, 95% confidence interval [0.7-2.7]). The regression model on the unmatched data showed similar results (OR = 2.08, 95% confidence interval [1.00-4.02] (Fig. 5). Models 2 and 4 which include an interaction term between sex and tattoo status showed higher effect compared to models 1 and 3 respectively. The matched data models 3 and 4 show similar yet more moderate effects when compared to unmatched data.

**Fig 5.**
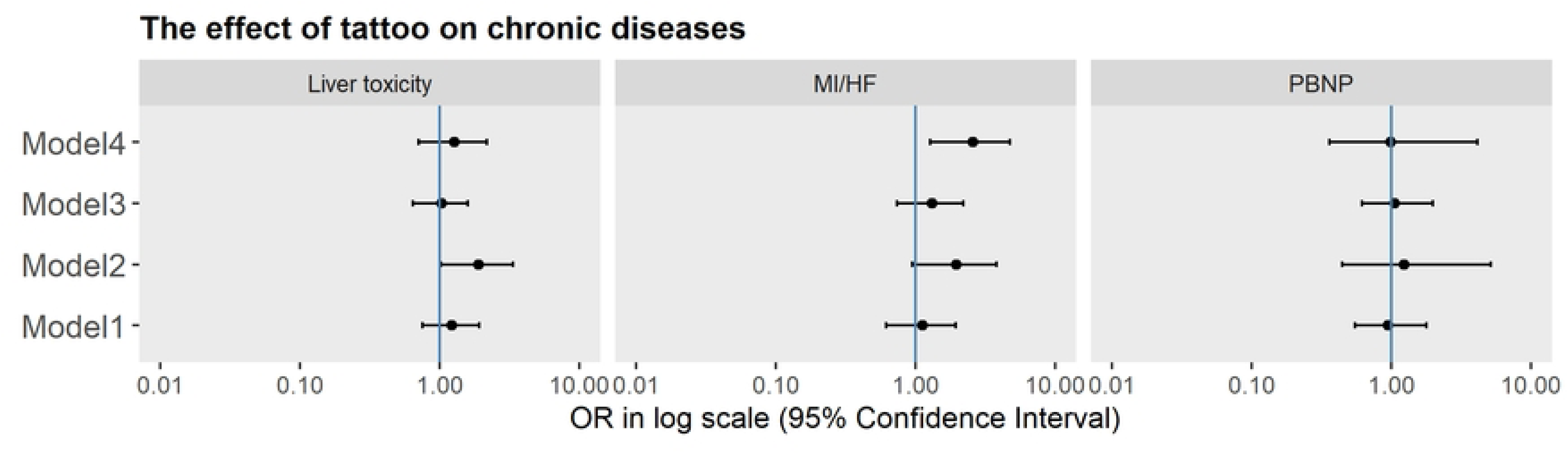
The effect of a tattoo on chronic diseases. Model 1 is a logistic regression on unmatched data and without interaction term between tattoo and sex. Model 2 is a logistic regression on unmatched data with an interaction term. Model 3 is a logistic regression on matched data without interaction term between tattoo and sex. Model 4 is a logistic regression on matched data with interaction term between tattoo and sex. MI/HF: Myocardial infarction and heart failure, PBNP: Pro-BNP.

Measured NT-proBNP values for tattooed and non-tattooed sub-cohorts at baseline examination are shown in Fig. S6. The prevalence of increased NT-proBNP levels is similar for tattooed/PMUed (13/202; 6.4%) and non-tattooed (251/3,892; 6.4%) individuals, but slightly different between men and women (men: tattooed: 3.5%, non-tattooed: 4.5%; women: tattooed: 8.2%, non-tattooed: 8.5%). The odds of increased NT-proBNP for tattooed/PMUed in matched data was 1.12, 95% confidence interval [0.41-4.65] in comparison to non-tattooed. The unmatched data showed similar results (OR = 1.23, 95% confidence interval [0.44-5.11]).

### Liver toxicity

Increased liver values indicative of hepatocellular damage were found in 11.3% (23/202) tattooed participants and 10% (390/3,897) non-tattooed controls (Fig. S7, Table S3). The size of tattoo and increased liver values did not show an association (Fig. 6a-d).

**Fig 6.**
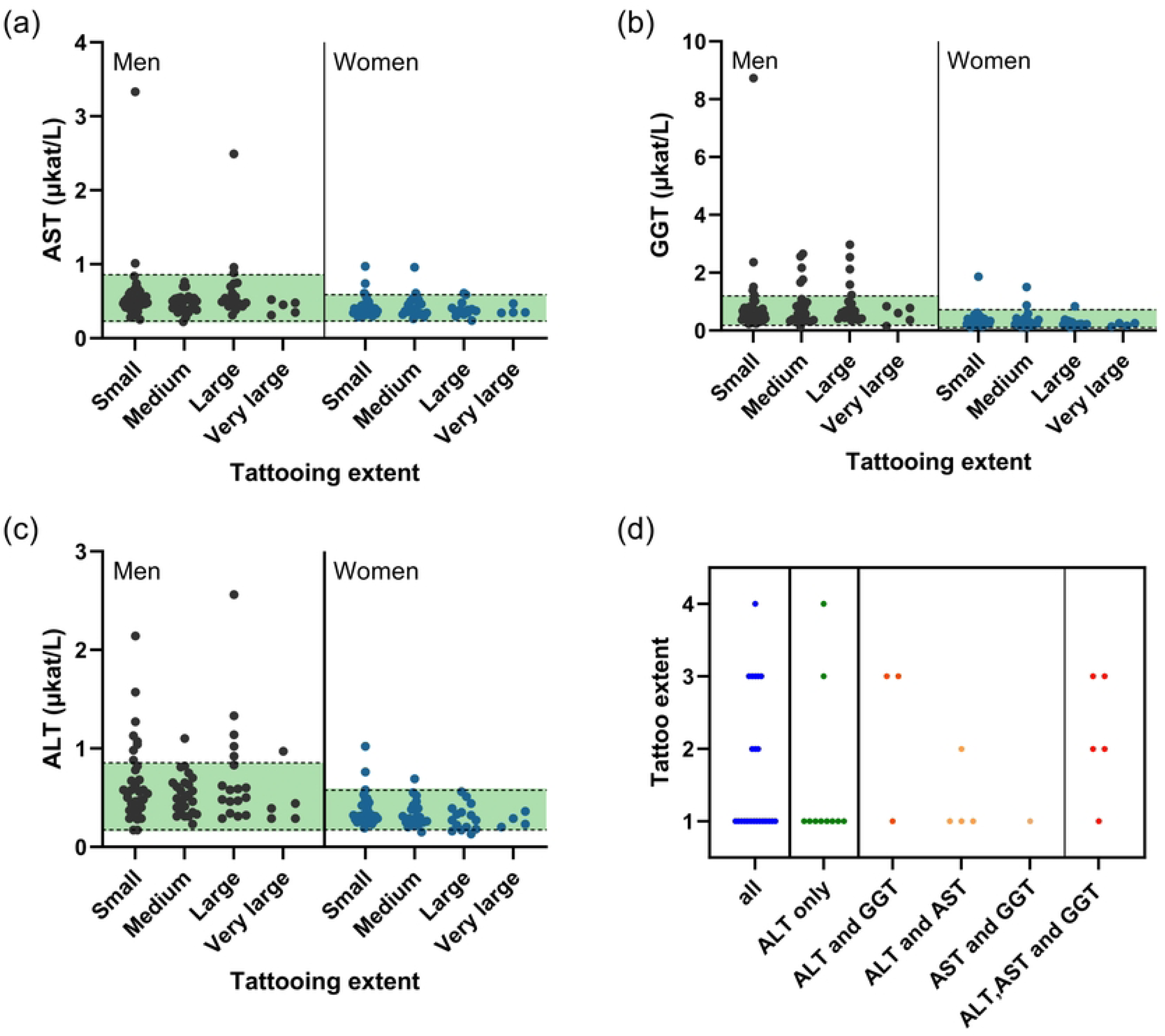
Correlation of tattooing extent and serum concentrations of diagnostic liver enzymes. A) ALT, B) AST and C) GGT. D) Tattooing extent (1 =small, 2 = medium, 3= large, 4 = very large) of all 23 participants with values indicative for a liver cell damage. ALT - Alanine aminotransaminase; AST - Aspartate aminotransaminase; GGT - γ-Glutamyltransferase.

The odds ratio for liver toxicity in matched data was 1.7, 95% confidence interval [0.95-2.96]. The unmatched data after adjusting for confounding showed similar results (OR = 1.9, 95% confidence interval [1.02-3.37]). In the sensitivity analysis we noticed a different effect among men and women and hence, added an interaction term between tattoo/PMU status and sex in our regression model. This is mainly due to a high risk ratio in men of 1.43 (95% CI: [0.86-2.37]), while being lower for women with 0.90 (95% CI: [0.41-1.96]) (Fig. 5).

### Cancer

Among the present tattooed/PMUed sub-cohort 16 cases of cancer occurred, as reported by the study participants. For 5/252 (2.0 %) of tattooed participants non-melanoma skin cancer including basal cell carcinoma (basaliom) and squamous cell carcinoma (spinaliom) were reported (Table S4). No melanoma cases were reported. Median age at self-reported diagnosis was reported to be 62 years (IQR 22 years). None of the participants stated a medical complication regarding their tattoo, indicating that the skin cancer did develop on other skin parts than the tattooed area. Other types of cancer are only reported in very few cases and therefore no further investigations were conducted.

### Sensitivity analysis

Results of the sensitivity analysis are shown in Fig. 7. We only were able to perform a male subgroup analysis for tattoos that took place before 1989 since the number of women who received a tattoo in this group was very small. Based on our model on unmatched data adjusted for confounding (age, sex, smoking status, socioeconomic status, BMI, and alcohol consumption) tattoos had an effect on liver toxicity in men before and after 1989. The difference between the effect of a tattoo on liver toxicity in men and women after 1989 remained consistent within the total sample.

**Fig 7.**
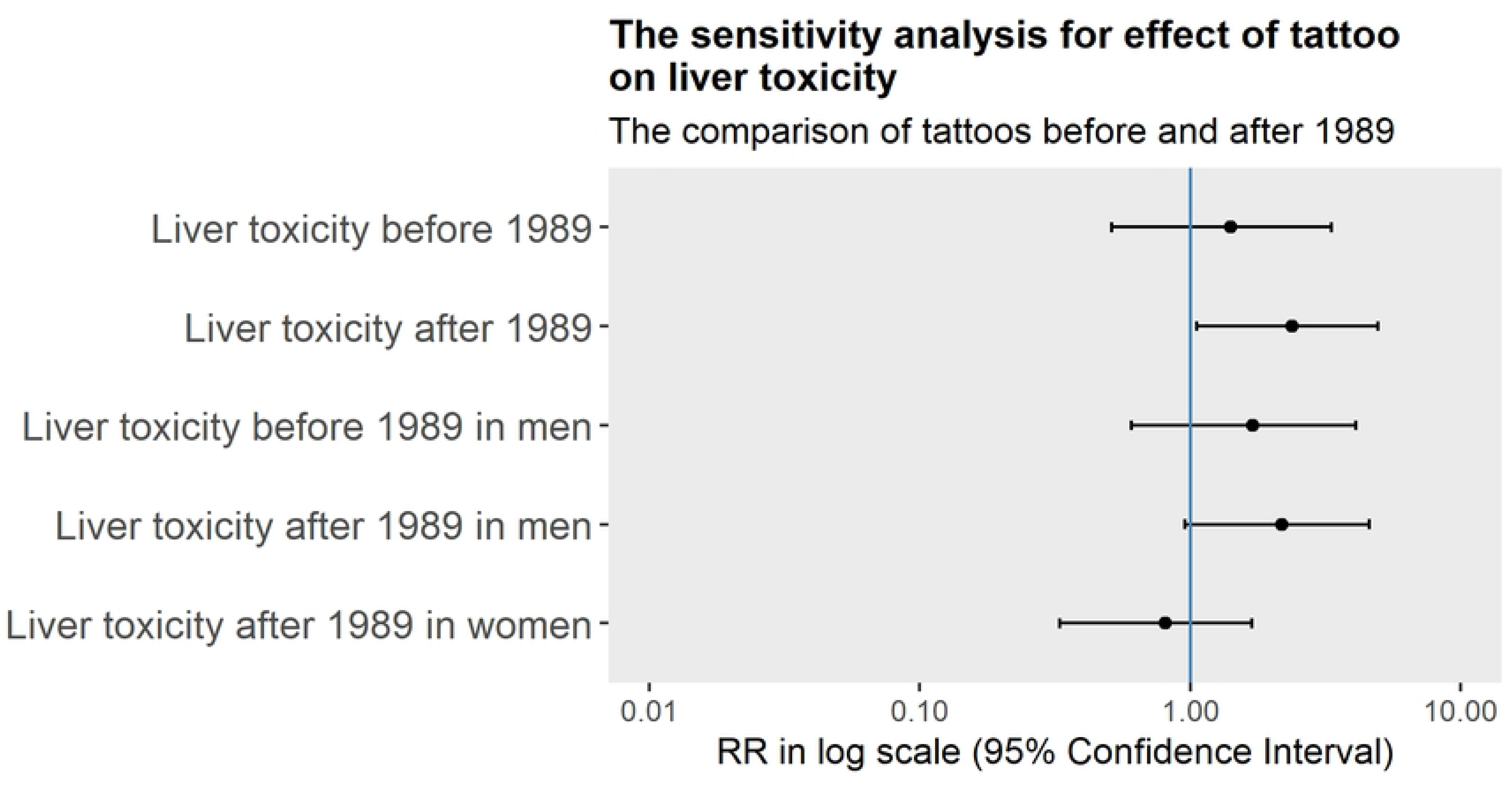
Sensitivity analysis to compare the liver effect of tattoos that took place before and after 1989.

## Discussion

The question of whether tattoos are associated with systemic health impairments is occupying researchers and regulatory authorities around the world. Despite the long-lasting practice of tattooing, robust epidemiological data is absent. In focus were rather case reports on cutaneous malignancies, indicating so far on coincidental causality between tattoos and the observed condition [36]. The very few case-control studies on tattoos and cancer indicate methodological inconsistencies [37, 38]. Only recently, Nielsen *et al.* demonstrated in a population-based case– control study that tattoos may comprise a risk factor for malignant lymphoma, indicating the urgent need for further comparable investigations [39]. With the present study, the relation of diverse systemic and chronic health effects such as cardiologic diseases, cancer and liver toxicity addressed within the LIFE-adult cohort and the presence and characteristics of tattoos and PMUs were investigated.

The tattooing incidence among each age group (Fig. 2e) is highest for 25-35 years with 14-18%, an observation in line with recent surveys showing higher prevalence within younger people [1–5, 40, 41]. In contrast, the PMUed subgroup has similar age distribution when compared to the overall cohort (median 71 years, IQR 14 years), and highest prevalence (4-5%) is found for participants between the ages of 60-75 years (Fig. 2d). 320/4,308 participants (7.4%) had either a tattoo (199, 4.7%) or a PMU (135, 3.1%) or both. This prevalence is lower in comparison to recent prevalence estimations among the general population of 10-20%. This might be due to the age distribution of the LIFE-cohort, which is not representative for the general population [29], with a profound over-representation in elderly generations, while tattooing is mostly popular among younger people.

### General medical issues related to tattoos/PMUs

The frequency of medical complications related to tattoos/PMUs within the present cohort is in general quite low with 5% (16/320) in comparison to findings of other studies reporting up to 57% [23, 42, 43]. However, in these studies the high frequency is mainly related to mild symptoms occurring immediately after receiving the tattoo/PMU and which not necessarily persist. Considering the long time passed since receiving the tattoo in the present study-cohort (Fig. 2c), it is possible that participants underestimated the symptoms.

Even though the frequency of pain, itch and swelling, most common medical complication named in relation to tattoos and PMUs in our study, is lower as in comparison to other studies, a high frequency of reactions to black tattoos/PMUs (7/10) is in line with results from Høgsberg *et al.* and Kluger [42, 43]. The tattooing extent among participants experiencing pain, itch or swelling was mainly small, indicating that the risk of occurrence of such complications is not necessarily related to the amount of ink injected.

Other complications named within the present study-cohort were herpes outbreak after receiving a PMU on the lips (n= 3, 0.9%), allergic tattoo/PMU reactions (n=2; 0.6%), scar formation (n=1, 0.3%) and a seldom and slight prickling at the site of the tattoo (n=1, 0.3%). Typically, herpes simplex virus type 1 (HSV-1) infection is the main cause of oral herpes [44]. It is known that triggers such as injuries, illness, stress or sun exposure can cause a reactivation of the virus. As the application of a PMU is coming along with small injuries at the site of needle penetration, it is plausible that these injuries might be the cause for the outbreak of the virus in the three reported cases.

The information gathered from the questionnaire does not allow to draw conclusions on the epitopes leading to the allergic reaction. However, in one case the participant stated that the initial red colour of the PMU changed to brown, indicating some extent of break-down of the pigments which may be correlated to the allergic reaction. There are indications that certain pigments, especially red colours, often cause allergic reactions [17, 24]. Some of the allergic reactions may occur immediately after tattooing and are then an indication of an existing allergy. Other reactions may occur months or even years after the tattooing, hence indicating that the allergy-causing component is probably formed by chemical degradation of the pigment or its metabolism within the body.

In both cases of reported skin reactions against metals, occurred for the first time after getting the tattoo or PMU, the participants did not indicate a tattoo related medical issue. However, this may indicate an initial sensitisation caused by allergenic impurities in the ink. Overall, within the present tattooed/PMUed cohort no indication for an increased risk on metal allergies was observed.

### Cardiologic effects

There are several case reports indicative for a risk for a bacterial endocarditis in correlation with receiving a tattoo [28, 45–51]. The reports are mainly among people with a history of congenial heart diseases indicative for a higher risk among sensitive subgroups. It is considered as a rare yet dangerous complication, as the endocarditis can lead to serious cardiac complications such as cardiac insufficiency or heart failure, requiring medical treatment and in most cases also surgical intervention [47]. Whether the risk for endocarditis is also given for people without congenial heart diseases or if any other coronary heart diseases such as MI or HF might be correlated to the presence of tattoos is to our knowledge not yet studied. We thus investigated cases of MI and HF among the present tattooed/PMUed cohort.

We found a positive odds ratio for cardiovascular diseases for tattooed participants. The matched and unmatched data showed similar results. However, the matched data showed slightly lower effect size, which may be attributed to a better control for confounding. While all cases of MI were men, in case of MI/HF positive risk difference origins from a risk ratio greater than 1 in men 1.96 [0.94-3.84], while in women risk ratio is much lower 0.5 [0.12-1.4]. Matched data analysis indicated a more moderate effect, yet suggesting a better control of confounding effects.

Unhealthy lifestyle habits such as smoking, overweight, and in particular, obesity, as well as related diseases such as diabetes and high blood pressure are main risk factors for MI as well as HF [52, 53]. In the majority of MI and HF cases (tattooed and non-tattooed) any or several of these risk factors where present (Table S2). The characteristics of cardiologic cases within tattooed and non-tattooed sub-cohorts are very similar with a high share of participants having known “life-style” risk factors for cardiologic complications such as smoking, overweight/obesity and diabetes mellitus. Average age of tattooed participants experiencing their first MI or MI/HF is slightly higher in comparison to the non-tattooed sub-cohort.

NT-proBNP is a physiologically inactive precursor peptide which is released together with the hormonally active and blood pressure reducing BNP by cardiac myocytes as a result of ventricular wall stress. Both peptides are therefore increased, dependent on the extent and duration of the ventricular dysfunction by HF, and are thus well-established parameters for diagnosing acute and chronic cardiac stress in adults [54, 55]. Only in one participant with HF, NT-proBNP levels were increased, which might be explained by the great time gap between determination of NT-proBNP at basic examination and the cardiac insufficiency event.

Among the reported cases of a tattoo-related infective endocarditis, which lead in many cases to HF, first symptoms were apparent closely after tattoo placement [28, 46]. Within the present cases of MI/HF, the majority of tattoos or PMUs have been present for a long time (>10 - 61 years). Thus, it is estimated unlikely that the cause for HF was a tattoo-related infective endocarditis. Overall, increased odds ratios among tattooed participants were found with a sex-specific difference.

### Liver toxicity

Liver diagnostics in form of determination of serum concentrations of the three enzymes AST, ALT and GGT was performed at baseline examination. An indication for toxic liver damage is established for increased ALT levels, increased GGT, AST and ALT, or AST and GGT combined [31–33].

Tattoo pigments were previously detected in Kupffer cells of the liver in mice [16]. Our data showed an increased risk for liver toxicity 1.7 [0.95-2.96]. The unmatched model showed a consistent effect. The matched data models (Fig. 5, Models 3 and 4) show similar but smaller effects compared to unmatched data, which most probably indicate a better adjustment for confounding effects. Even if no robust conclusion can be drawn due to a low sample size and possible residual confounding, the results suggest that liver toxicity of tattooing pigments should be further investigated.

### Cancer

So far only few case-control studies investigated cancer risk associated to the presence of tattoos [37, 38]. Barton *et al.* studied the risk for basal cell carcinomas at the site of a tattoo in 156 cases [38]. Their results support the notion that tattoos could enhance the risk of an early onset of basal cell carcinomas, however, these findings are only preliminary due to the small number of cases investigated. In addition, some individual cases on malignancies and benign tumours at the side of the tattooed area are reported, however in none of these cases tumour development could be correlated with certainty to the tattoo [36, 56, 57]. Warner *et al.* investigated the correlation between tattoos and lymphatic cancer, as it is known that tattoo pigments are transported to local lymph nodes and thus cancer development at these sites would be biologically plausible [37]. The authors did not identify any significant associations between tattoos and the risk of developing lymphatic tumours. In the publication by Nielsen *et al.*, 2024, the influence of tattoos on the development of malignant lymphomas in the Swedish population was investigated using a population-based retrospective case-control study [39]. The incidence rate for lymphoma in tattooed participants was increased by 21% (IRR 1.21) compared to the control group. However, uncertainties associated with the subsequent analysis of the influence of exposure remain.

Among the present tattooed sub-cohort 5 cases of non-melanoma skin cancer including basal cell carcinoma (basaliom) and squamous cell carcinoma (spinaliom) were reported (Table S4). None of the participants stated a medical complication regarding their tattoo, indicating that the skin cancer did develop on other skin parts than the tattooed area. In general, non-melanoma skin cancer is the most common type of cancer overall [58] with UV exposure and age as highest risk factors for its development [59]. As such it is expected that non-melanoma skin cancer has a relatively high frequency among our tattooed cohort. As the overall number of cases within our cohorts are low, no further analysis was performed.

Our study had some limitations which we think might affect the results. First, the tattoo/PMU data as well as the health outcomes were self-reported (questionnaire). The characteristics of tattoo exposure, such as time point of tattooing, size, colour, are estimated to being less significant when collected via self-report. Second, the number of confounding factors that could be controlled for were limited. As shown in the DAGs (Fig. S2, Fig. S3), a residual confounding may be present. This might also explain the difference in effect observed among men and women. Modern computational methods were used and compared to classical logistic regression to show the consistency of the results when analysed via different methods. However, the matched data was found to be more efficient to account for confounding and result in odd ratios which better represent the effects. Third, the study participants were all residents in the East of Germany and results should be cautiously interpreted for other populations.

## Conclusion

In this study a population bearing a tattoo or a PMU was thoroughly characterised. Despite the low prevalence, it permitted the analysis of systemic health effects, benefited from the long exposure of over 10 years. Tendency for elevated cardiologic effects as well as liver toxicity could be observed, but needs further confirmation. Since the composition of the applied tattoo inks is unclear, the data do not allow a conclusion of such risks by current tattoo practices. Our results highlight the urgent need for the elucidation of chronic effects of tattoo inks, which could underpin biological plausibility of respective findings. Such studied should be conducted with thoroughly characterised (analysed) tattoo inks, since small changes in chemical composition or impurity profile may alter toxicological effects [60]. Moreover, further epidemiologic studies among larger cohorts are required, which could mitigate or strengthen the findings of this study.

## Data Availability

The raw data for this study are stored on the servers of the Leipzig Research Center for Civilization Diseases (LIFE), University of Leipzig, Germany. The data are clearly stored under the project PV-Wirkner-2021-602-10. It can be requested in accordance with the LIFE Research Center's guidelines on the release of data to third parties. Please address inquiries regarding these procedures to the LIFE Data Management.

## Supporting information

**S1 Questionnaire. Tattoo-Questionnaire (original in German).**

**S1 Fig. Unadjusted Directed Acyclic Graph (DAG) of the tattoo effect on chronic health outcomes.**

**S2 Fig. Adjusted DAG of the tattoo effect on chronic health outcomes after adjusting for the parameters in the model.**

**S3 Fig. Tattoo/PMU colours. Colour distributions of tattoos and PMUs among men and women.**

**S4 Fig. Tattoo location and size.** Participants were asked to state the location and respective size of their tattoos. Thereby, the sizes of tattoos were estimated according to four categories (<5, 5-10, 10-20, >20 cm). These may be interpreted as proximal length of the tattooed area. Thus, the area of the tattooed body surface was calculated as a square on the basis of the given sizes as length of one side of the square. The four categories translate into areas as follows: <25 cm² (average 13 cm²), 25-100 cm² (average 62.5 cm²), 100-400 cm² (average 250 cm²), > 400 cm². It is to note that this is a very rough estimate of tattoo size and area. It results from a self-assessment of the participants estimating their tattoo size and neither different shapes of tattoos were accounted for, nor a differentiation between a fully, or partially coloured area or outlines only was made. Thus, the area of the tattooed body surface is probably overestimated in general. For body tattoos, most popular location was the arm with overall 170 counts (51%), thereof 84 tattoos (25 %) were at the upper arm and 86 (26 %) were at the forearm including wrist. This is followed by back (55 counts, 17%) and torso (42 counts 12%), in both cases the upper regions were more common than below the waist (upper back 43 counts (13%) and upper torso 27 counts (8%)). PMUs were almost exclusively located in the face (126 counts). 8 participants stated to have additionally a tattoo. 1 participant indicated to have a PMU at the head, but other than in the face. No other body regions were named in correlation to a PMU. The highest share of very large (> 400 cm²) and large (100-400 cm²) tattoos was observed at the front thighs, followed by upper torso, upper back, upper arm and calf (Figure S5). Concordant, the biggest share on small (< 25 cm²) to medium (25-100 cm²) size tattoos (or PMUs) are at body regions of smaller area such as face, neck, hands and feet. On the arm, tattoos of medium (25-100 cm²) to large (100-400 cm²) sizes were predominant.

**S1 Table. Medical issues related to tattoos and PMUs (self-estimation of participants).**

**S2 Table. Characteristics of tattooed/PMUed participants in comparison to non-tattooed/PMUed who experienced myocardial infarction (MI), and heart failure (HF)/heart insufficiency (HI).**

**S5 Fig. Heart biomarkers.** NT-proBNP at baseline. Norm values for NT-proBNP: from 18 years <115 pg/mL, from 45 years <172 pg/mL, from 55 years < 263 pg/mL, from 65 years < 349 pg/mL, from 75 years <738 pg/mL (Universitätsklinikum Leipzig).

**S6 Fig. Liver biomarkers.** Distribution of measured enzyme concentrations in serum: Overall, 44 out of 202 tattooed/PMUed participants had an elevation in at least one of the liver values (Figure S7). However, 31 participants had only one value elevated, thereof 6 participants have only AST and 15 only GGT increased, which is assessed as less relevant regarding liver toxicity.

**S3 Table. Characteristics of tattooed/PMUed and non-tattooed/PMUed participants with liver diagnostic values indicative for a hepatocellular damage.**

**S4 Table. Tattooed participants with a diagnosed non-melanoma skin cancer (results from basic and follow-up examinations combined).** For the evaluation, only participants from which tattoos were present before answering the questionnaires and cancer diagnosis were included.

## Acknowledgements

The authors would like to thank Dr. Milena Förster for her consultation and productive discussions to the earlier drafts of the manuscript. A warm thanks also to Dr. Agnes Schulte for her support und supervision.

## Author Contributions

Conceptualization: Loryn E. Theune, Narges Ghoreishi and Michael Giulbudagian. Data curation: Christoph Engel, Kerstin Wirkner, Ronny Baber. Writing: Loryn E. Theune, Narges Ghoreishi, Christine Müller-Graf, Peter Laux, Andreas Luch, Michael Giulbudagian. All authors commented on previous versions of the manuscript. All authors read and approved the final manuscript.

## Ethics approval

The responsible ethics board at the Medical Faculty of the University of Leipzig approved the study (reference: LIFE Adult - 263-2009-14122009).

## Consent to participate

All participants provided written informed consent to participate prior to participation.

